# THE DYNAMIC AFFECT RECOGNITION TEST: CONSTRUCTION AND VALIDATION IN NEURODEGENERATIVE SYNDROMES

**DOI:** 10.1101/2024.12.23.24319565

**Authors:** Katherine P. Rankin, Hulya Ulugut, Anneliese Radke, Scott Grossman, Pardis Poorzand, Tal Shany-Ur, Joel H. Kramer, Katherine L. Possin, Virginia E. Sturm, Maria Luisa Gorno Tempini, Bruce L. Miller

## Abstract

**Learning objective:** To validate a novel video-based emotion identification measure in persons with neurodegeneration and show correspondence to emotion-relevant brain systems

**Background:** Given advances in disease-modifying therapies for dementia, the dementia field needs objective, practical behavioral assessment tools for patient trial selection and monitoring. The Dynamic Affect Recognition Test (DART) was designed to remedy limitations of instruments typically used to measure emotion identification deficits in persons with dementia (PWD).

**Method:** Participants included 372 individuals, including 257 early stage PWD (Clinical Dementia Rating ≤1, Mini-Mental State Examination ≥20; 66 behavioral variant frontotemporal dementia [bvFTD], 27 semantic variant primary progressive aphasia [svPPA], 23 semantic bvFTD [sbvFTD], 33 non-fluent PPA [nfvPPA], 26 progressive supranuclear palsy [PSP], 28 corticobasal syndrome [CBS], 42 Alzheimer’s disease [AD], 12 logopenic variant PPA [lvPPA]), and 115 healthy controls (HC), watched 12 15-second videos of an actor expressing a basic emotion (happy, surprised, sad, angry, fearful, disgusted) via congruent facial/vocal/postural cues, with semantically neutral scripts. Participants selected the emotion from a randomized visual array. Voxel-based morphometry (VBM) analysis was performed to show brain structure correlates of DART, controlling for non-emotional naming ability (Boston Naming Test, BNT).

**Results:** DART performance was worse in PWD than older HC (p<0.001), with the lowest scores observed in the sbvFTD group. A DART 10 cut-off score differentiates PWD from HC with a 90% sensitivity and 49% specificity (AUC=82%). A DART 9/12 score yielded 93% sensitivity/67% specificity (AUC=87%) for discriminating social cognition disorders from HC, while a 7/12 score differentiated sbvFTD from HC with 100% sensitivity/93% specificity (AUC=97%). VBM showed poorer DART performance significantly predicts focal brain volume loss in right-sided emotion processing areas including insula, temporal pole, caudate, superior frontal gyrus and supplementary motor cortex (pFWE<0.05).

**Conclusions:** The DART is a brief, psychometrically robust video-based test of emotion reading (i) designed to be practically useful in realistic assessment settings, (ii) effectively reveals emotion identification impairments in PWD, (iii) shows specificity for identifying PWD exhibiting real-life SCDs (i.e. bvFTD, svPPA, sbvFTD), (iv) corresponds to the expected structural anatomy of emotion reading, and (v) is freely available to researchers and clinicians.

## INTRODUCTION

Some individuals experience a reduced capacity to accurately read emotional expressions as a result of neurodegenerative disease. It is well-established that individuals with behavioral variant frontotemporal dementia syndrome (bvFTD)(1) show disproportionately impaired emotion processing in their daily lives, particularly compared to those with Alzheimer’s disease syndrome (AD) (2) and to some of the primary progressive aphasias (PPA), (3) including the non-fluent variant (nfvPPA) and the logopenic variant (lvPPA) syndromes, in which emotion reading is comparatively spared (4,5). The semantic variant syndrome (svPPA), however, is associated with substantial emotion reading deficits.(5) More recently, there has been a movement in the dementia field to separate these “temporal variant” patients into two categories, classifying patients with left temporal-predominant disease as svPPA, but establishing a distinct category for patients with predominantly right anterior temporal lobe (RATL) atrophy (6,7). These patients, which some have called semantic bvFTD (sbvFTD),(8) have early socioemotional deficits and show particularly severe problems with emotion recognition and naming, the mechanism of which seems to involve semantic loss for emotions (5,9–11). Both the svPPA and sbvFTD variants predominantly show dysfunction in the semantic appraisal network of the brain (6,8,12–14). In, bvFTD degeneration originates in the salience network (SN) (15), which implicates this network in these individuals’ characteristic empathy loss. Together, this patient evidence suggests that altered emotion reading is mechanistically complex in neurodegenerative disease, and may involve both salience and semantic appraisal networks.

Using objective, practical, time effective, and culturally sensitive neuropsychological assessment tools is pivotal for accurate patient selection and monitoring of symptom progression in clinical trials, which is increasingly urgent given substantial advances in disease modifying therapies for neurodegeneration. Since 1976 when Ekman and Friesen published their rigorous standards for representing facial emotion expressions (16), the dominant approach to testing patients’ emotion recognition in both research and clinical settings has been to use still photographs of emotional faces as stimuli. Examinees view a static image and may be asked to spontaneously name the emotion depicted, choose the emotion from among a multiple-choice array, or match the emotional face with another emotional face. However, these static face stimuli leave out many important aspects of real-world emotion expression, such as movement dynamics, vocal prosody, body posture, and gestures, all of which are part of the naturalistic interpersonal signaling normally used to identify another person’s emotional state. Several groups have called into question the ecological validity of using static facial expressions to identify real-life emotion recognition impairments (17–24). The main arguments have been that static photographs may not be “as emotionally rich” as the other measures such as prosody, (17,21) or “facial photographs lack important dynamic information the dementia patient needs to more accurately interpret facial expressions”(22). Particularly when examining patients with cognitive deficits due to neurodegenerative disease, using test stimuli artificially limited to only the visual channel of what is naturally a multi-modal and dynamic display makes emotion recognition tests abnormally difficult, and thus results may not match an individual’s real-life emotion reading capacity (20). Recent meta-analyses of studies of emotion reading in individuals with neurodegenerative disease reveal substantial variability in results across studies, likely influenced by differences across tasks (4,25,26). Furthermore, one study has found that tasks depicting real-life scenarios had greater sensitivity with bvFTD patients than other neuropsychological measures. (23) Overall, there is increasing consensus that more ecologically realistic, video-based stimuli incorporating both visual information (not only facial expressions, but also direction of gaze, posture, gestures), and auditory signals (prosody, volume of speech) are necessary to provide a more neuropsychologically valid reflection of emotion reading capacity in patients with cognitive deficits.

One test comprising dynamic videos for emotion testing that has been successfully employed in patients with neurodegeneration is the Emotion Evaluation Test (EET) subtest of The Awareness of Social Inference Test (TASIT) battery. (27) It asks patients to label realistic video displays of seven emotional expressions (happy, surprised, sad, anxious, angry, disgusted, and neutral) that are conveyed using a semantically neutral script (i.e., so that the examinee must determine the emotion from paralinguistic cues, not from spoken content). However, some characteristics of the TASIT-EET can limit its utility in cognitively impaired patient samples. It is a commercially-distributed test, which prices it out of range for use in many clinical and research settings. Further, videos depict multiple actors onscreen, requiring the examinee to track and retain identity information (including name) in order to identify whose emotion should be targeted. The actors have Australian accents, which could reduce comprehension by patients in other English-speaking contexts. Finally, the test is administered by an examiner using pencil and paper answer sheets, which adds to task complexity and reduces access.

In response to these limitations, we developed the Dynamic Affect Recognition Test (DART) specifically for measuring emotion identification deficits in cognitively impaired patients. The DART is a digitally administered and freely-available 12-item emotion labeling task with 6 emotions (excluding neutral). Each video scene is simplified, depicting a single actor in front of a minimal background with few environmental distractions, and it uses racially and ethnically diverse actors with American accents. Here we describe the approach to construction, construct validity, patterns of performance across neurodegenerative disease syndromes, and corresponding brain anatomy of the DART. We hypothesized that DART performance would show divergent, clinically predictable patterns across neurodegenerative patient groups, and would correlate with focal atrophy in brain systems known to mediate emotion reading and naming, both of which would support its criterion validity. Finally, we provide more information about the effectiveness of the DART in making differential diagnostic decisions and suggest cut- off scores for making particular discriminations among neurodegenerative syndromes.

## METHODS

### Participants

All participants were diagnosed on the basis of a comprehensive multidisciplinary evaluation, and included a total of 372 individuals: 257 persons with dementia (PWD) and 115 healthy controls (HC). In the PWD group, 66 participants met consensus diagnostic criteria for bvFTD, (1) 27 svPPA, (3) 23 semantic bvFTD (sbvFTD), (8) 33 nfvPPA, (3) 12 lvPPA, (3) 26 with progressive supranuclear palsy syndrome (PSPS), (28) 28 corticobasal syndrome (CBS), (29) and 42 individuals with biomarker-confirmed AD.(2) To ensure a very early stage of disease severity across syndromes, participants were included if they had a Clinical Dementia Rating Scale (CDR) (30) Global Score ≤1, CDR plus National Alzheimer’s Coordinating Center (NACC) behavior and language domains (FTLD) Sum of Boxes (31) scores ≤8, and Mini Mental State Examination (MMSE) (32) score ≥20.

Healthy controls were English-speaking predominantly well-educated individuals from the San Francisco Bay Area who had undergone complete neurologic, neuropsychological, and MRI evaluation and had normal MRI and neurologic exam and no cognitive or functional deficits. The majority of the participants were of European ancestry (89.3%), with 5.4% reporting Asian ancestry, 3.0% reporting Latino/Hispanic ancestry, and 2.4% mixed/other ancestry. While HCs of all ages were analyzed to derive normative characteristics for the DART (Table 2), only the subset of older HC individuals (OHC; age range 46-90; N=71) were used in statistical comparisons with the PWD cohort.

### Procedures

#### The Dynamic Affect Recognition Test (DART)

<u>Test construction:</u> The DART was designed to provide a measurement of emotion reading capacity that is valid even in individuals with other non-emotional cognitive deficits in the context of a rapid dementia evaluation, thus primary requirements for the test were a) the test should use video stimuli, with actors expressing emotions via realistic visual and vocal/auditory cues including upper body and head movements and posture, b) it needed to be as brief as possible to reduce participant cognitive fatigue and to make it more likely that the test would be administered in time-constrained clinical research settings, c) at minimum it should include the 6 primary and most universally-recognized emotion expressions (happy, surprised, sad, angry, frightened, and disgusted), with expressions validated based on the Facial Affect Coding System (FACS),(33) d) stimuli should be as simple as possible, including only one actor on screen with a non-descript background setting, e) actors should verbalize scripted lines, either alone or in dialogue with an off-screen interlocutor, in order to make their emotional expressions as realistic as possible; however, to ensure that emotion reading is based only on paralinguistic (i.e. facial, prosodic, postural, gestural, movement) cues rather than on content inference, the scripts should have only emotionally ambiguous content, i.e. words that could be stated with any of multiple different underlying emotional states, making the content of the spoken words irrelevant to the task of determining what emotion the speaker is expressing, and f) the task should be available via non-commercial means of distribution, i.e. freely available for use by qualified clinical researchers and clinicians.

Scripts for the task were written in English and edited collaboratively by the research team. Actors fluent in American English who were of diverse ancestry, age, and sex were selected. Each actor was given a single item script to learn, but was filmed expressing that script with all six emotions, with films lasting ∼10-15 seconds each. The videos underwent consensus evaluation by the team to identify the two best expressions of each of the six emotions, and those videos underwent additional Facial Affect Coding System (FACS) coding by a blinded rater to confirm that actors were showing key facial action units associated with the target emotion, and were not showing significant additional off-target emotion expressions. Final face validity for each item was evaluated after the first twenty healthy controls had performed the task, and confirmed that for every item >85% of HCs agreed on the target emotion depicted by the item and obtained a correct response. The task was constructed to contain an easy example item (happy) plus 12 test items (pseudo-randomly representing two items for each of the six basic emotions). After presentation of each video, participants are shown a screen with all six emotion names, with positions pseudo-randomized across items, and provided with the instruction to select the emotion that the person in the video just showed (i.e. touch the word on a tablet, or select it via mouse click on a computer). Participants can change their response until satisfied, then select a “next” button on the screen to confirm their final answer and move on to the next item. Full administration of the task can take from 3-5 minutes, depending on self-paced speed.

<u>Administration.</u> The DART was built to into two presentation platforms to facilitate more widespread use: 1) the TabCAT tablet-based cognitive assessment platform, designed and hosted at UCSF (34), and available for free download for iPadOS from the Apple Store, and 2) a generic version of the task was made available that is suitable for users to upload test materials into any online survey administration program. For online administration at UCSF, the Qualtrics survey platform was used (https://www.qualtrics.com/). While the DART can be independently self-administered, for the purposes of this study it was performed in the presence of a trained psychometrist who was available to ensure participants understood and complied with test instructions. Participants had the option to respond to items either by pointing/touching their answer on the screen or, in cases where cognitive or motor deficits precluded autonomous self-administration, by verbally stating their response to the examiner, who then recorded it.

#### Additional Cognitive and Functional Measures

Neuropsychological testing data from participants’ diagnostic evaluations were also included in this study. These included the Mini Mental Status Examination (MMSE) to gauge global cognitive functioning, (32) and a 15-item abbreviated version of the Boston Naming Test (BNT) (to evaluate capacity for confrontation naming of objects). (35) Disease severity was evaluated using the Clinical Dementia Rating (CDR) Scale Global Score (30) as well as the CDR plus NACC FTLD Global and CDR plus NACC FTLD Sum of Boxes scores (31) (for more information also visit https://naccdata.org/data-collection/forms-documentation/uds-3). All neurocognitive and disease severity data were collected on average within a week before or after completing the DART (with a maximum time window set at 90 days).

#### Structural MRI Acquisition and Preprocessing

All participants underwent structural T1-weighted images acquired on 3T-scanners (Siemens Trio and Siemens Prisma). A T1-weighted 3D magnetization prepared rapid gradient echo (MPRAGE) sequence was used, with acquisition parameters as follows: 160 sagittal slices, 1- mm thick, skip = 0 mm; repetition time = 2300 ms; echo time = 2.98 ms; flip angle = 9°; field of view = 240 × 256mm2; voxel size = 1mm3; matrix size = 256 × 256.

Structural T1-weighted images were preprocessed for voxel-based morphometry (VBM) analysis via the UCSF Brainsight system (36). The images were visually inspected for artifacts, and underwent bias-correction, segmentation into tissue compartments, and spatial normalization using a single generative model with the standard SPM12 toolbox parameters. The default tissue probability maps for grey matter, white matter, cerebrospinal fluid, and all other voxels from SPM12 (TPM.nii) were used (37). To optimize intersubject registration, each participant’s image was warped to a template derived from 300 confirmed neurologically healthy older adults (ages 44-86, M±SD: 67.2±7.3; 113 males, 186 females) scanned with one of three magnet strengths (1.5T, 3T, 4T), using affine and nonlinear transformations with the help of the diffeomorphic anatomical registration through exponentiated lie algebra (DARTEL) method, with standard implementation in SPM12 (37). Total volume of each tissue compartment was calculated by applying the modulated, warped and segmented masks for gray matter, white matter, and CSF to the corresponding MWS probability map for that individual, and the total intracranial volume (TIV) was derived by summing the three volumes. The spatially normalized, segmented, and modulated gray matter images were smoothed with an 8-mm FWHM isotropic Gaussian kernel for use in VBM analysis.

### Statistical Analysis

Analysis of diagnostic group differences on potential confounds (age, sex, education, MMSE score) was conducted to identify covariates that were then included in later analyses. Also, regression diagnostics were performed on the DART total score to identify inappropriately influential data points, as well as to examine the normality, heteroscedasticity, and multicollinearity of residuals. Differences among all diagnostic groups were explored for the CDR, global CDR® plus NACC FTLD score, CDR® plus NACC FTLD Sum of Boxes, MMSE total score, BNT total score, and DART total score using the analysis of variance (ANOVA) test. Significant differences between diagnostic groups and controls were identified using Dunnett– Hsu post-hoc tests, whereas differences among diagnostic groups were examined using Tukey post-hoc tests (Table 1).

**Table 1:** DART normative reference table, neurologically healthy controls only.

| **Control Cohort** | **Age 19-39**<br>**(N=41)** |  | **Age 40-69**<br>**(N=41)** |  | **Age 70-90**<br>**(N=33)** |  |  | **Male**<br>**(n=50)** |  | **Female**<br>**(n=65)** |  |  |
| --- | --- | --- | --- | --- | --- | --- | --- | --- | --- | --- | --- | --- |
|  | **Mean** | **SD** | **Mean** | **SD** | **Mean** | **SD** | **p-value** | **Mean** | **SD** | **Mean** | **SD** | **p-value** |
| **Sex (male/female)** | 17/24 | - | 16/25 | - | 17/16 | - | n.s. | 50 | - | 65 | - | n.s. |
| **Education** | 16.1 | 3.4 | 17.6 | 2.1 | 17.9 | 2.6 | n.s. | 17.3 | 2.8 | 17.2 | 2.8 | n.s. |
| **DART Total score** | 10.8 | 0.2 | 10.6 | 0.2 | 9.7 | 0.2 | 0.01 | 10.1 | 1.1 | 10.6 | 1.2 | 0.02 |

**Table 1:**
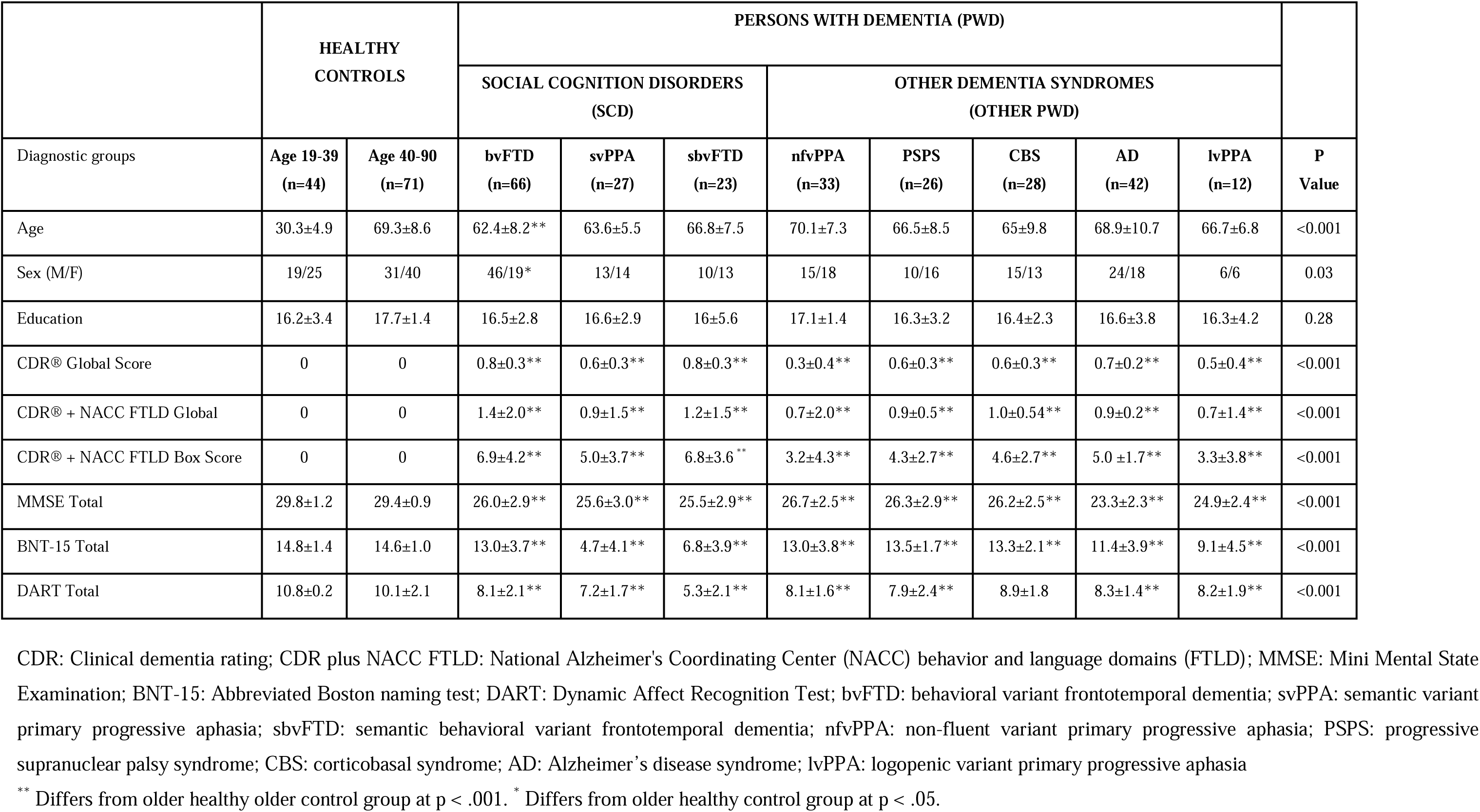
Clinical cohort demographics and clinical characteristics.

### DART Validation Analyses

#### Construct validity

Convergent validity of the DART was examined by determining if it produces results consistent with known patterns of emotion recognition symptoms in patients. Diagnostic group differences on DART performance (total score) were evaluated using a general linear model (SAS PROC GLM), adjusting for age and sex as confounds, with Dunnett-Hsu post-hoc tests comparing each diagnostic group to the NC group.

<u>Derivation of cutoff scores for differential diagnosis.</u> To provide more information about the effectiveness of the DART in making differential diagnostic discriminations among PWD, as well as to determine appropriate score cutoffs to be used in those discriminations, receiver-operating characteristic (ROC) analyses were performed. Area under the curve (AUC) derived from to test the accuracy of the DART in PWD versus OHCs, the subset of PWD with social cognition deficits, i.e. SCD (bvFTD+ svPPA+ sbvFTD) versus HCs, sbvFTD versus OHCs, SCD versus other PWD and sbvFTD versus other PWD. Sensitivity and specificity values were calculated for each score in order to determine appropriate cutoff scores for these pairwise discriminations. Sensitivity and specificity thresholds for patient classification were derived from these ROC plots and 2x2 contingency table analyses in order to determine appropriate cutoff scores for these pairwise discriminations. A K-fold cross validation technique was used to improve our methodology. This method guarantees that the score of our model does not depend on the way we picked the train and test set. The data set is divided into k number of subsets and the holdout method is repeated k number of times (38). We calculated sensitivity (also called true positive rate) as the chance that DART score will accurately identify the patients belonging to a diagnostic group, and specificity (also called true negative rate) as the chance a patient that does not belong to a group is accurately excluded (e.g., a healthy control will not be identified as a dementia patient).

<u>Concurrent (i.e. predictive) validity</u> of the DART was examined by identifying whether DART score corresponded with volume in expected neuroanatomic regions, i.e predominantly regions known to be associated with emotion reading. We performed a whole brain analysis of atrophy patterns corresponding to individual differences in DART performance, including all PWD and OHC participants (N=328). To do this, we performed a voxel-based morphometry (VBM) analysis of gray matter maps using a covariates-only design matrix with DART score as the primary correlation, with the following covariates: age, sex, and total intracranial volume (TIV) as nuisance covariates [1 0 0 0 . . .]. To further evaluate the degree to which DART score predicted brain volume specific to emotion naming, distinct from general confrontation naming ability, DART score was evaluated against gray matter volume controlling for total score on the BNT, with the same nuisance covariates. To determine the threshold for family-wise error correction, maximum t-values of the imaging data compared to behavioral data were re-sampled using 1000 permutations of the error distribution in a Monte Carlo approach. Maximum t-values for each permutation were used to create a custom error distribution, and the t-value at the 95th percentile of this distribution was taken as the custom cut-off threshold, rendering t-values on or above this cut-off significant at a family-wise error corrected level of p<0.05 (39).

## Ethical approval

This research was subject to approval by the UCSF Committee for Human Resources Independent Review Board. In all cases, informed consent was gained from the participant or the primary caregiver.

## Data availability

Individual-level data are available in the access-controlled FAIR Alzheimer’s Disease Data Initiative AD Workbench repository at https://fair.addi.ad-datainitiative.org.

## RESULTS

### Demographic and Clinical Characteristics

<u>Healthy control cohort:</u> Normative reference tables including demographic characteristics and DART scores subdivided by age and sex groups are found in Table 1. The healthy control cohort was highly educated on average, thus additional subgroupings by education level could not be performed.

<u>Full clinical cohort:</u> Demographic and clinical characteristics of all healthy controls and PWD are found in Table 2. Statistical comparisons across groups were performed using only the N=71 older healthy controls (OHC), excluding younger controls from further analysis. Few statistical differences were observed, though on average the bvFTD group was significantly younger than OHC, and were more likely to be male. CDR and MMSE scores, representing disease severity across PWD groups, showed that all patients were at very early disease stages, reflecting our strict inclusion criteria.

### Accuracy of DART for identifying diagnostic group differences in PWD

Compared to OHCs, all PWD groups performed significantly worse on the DART (p< 0.001), with the exception of the CBS group (Table 2). The lowest DART scores were observed in sbvFTD, with a mean DART score of 5.3±2.1, and a significant difference was observed in sbvFTD group compared to all other dementia syndrome groups after Tukey-Kramer post-hoc testing controlling for age and sex (p<0.05). (Figure 1).

**Figure 1:**
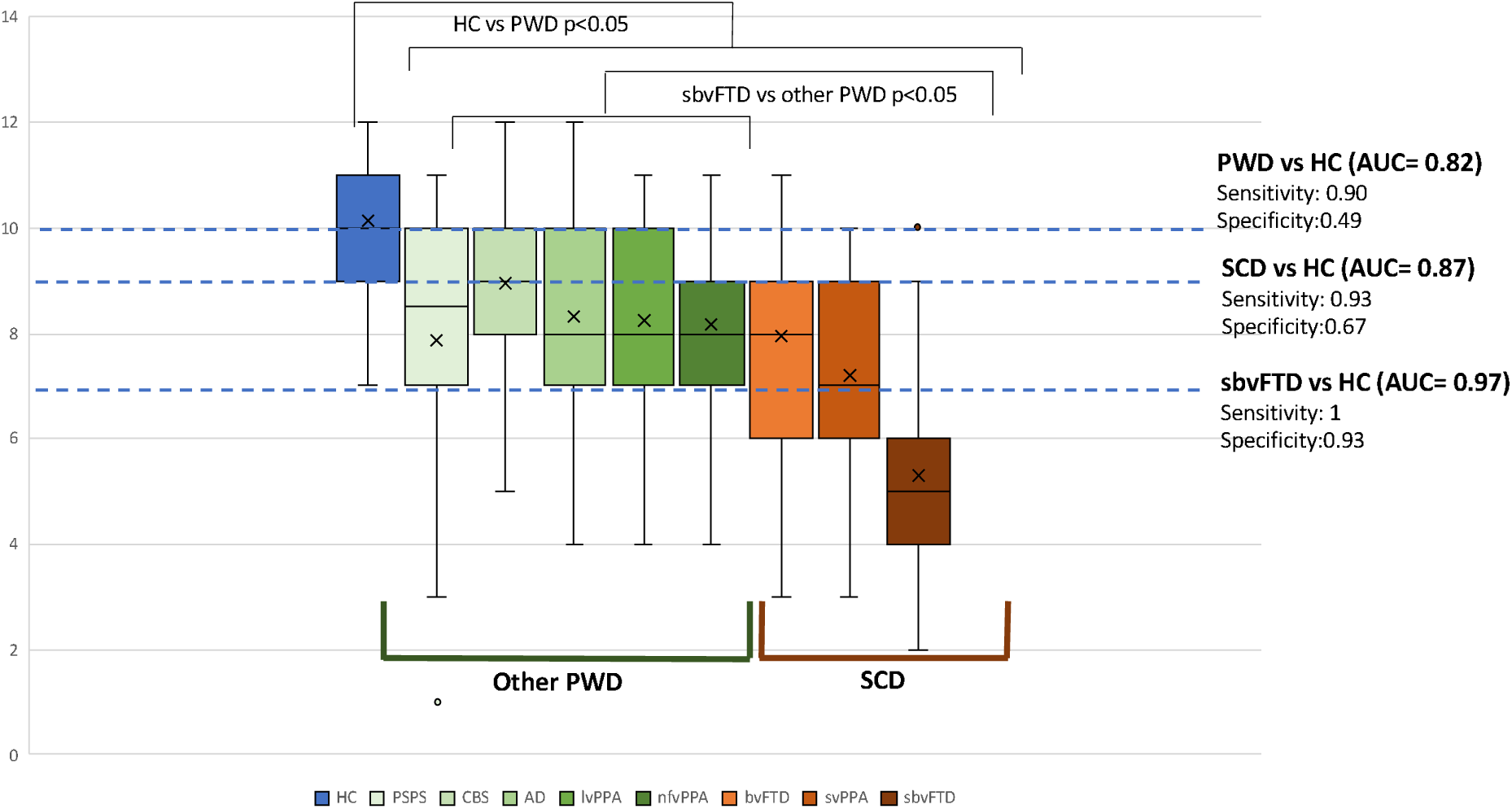
Total DART scores across diagnostic groups and cut-off values. PWD: patients with dementia, SCD; social cognition disorders; AUC: area under the curve; OHC: older healthy controls; CBS: corticobasal syndrome; AD: Alzheimer’s disease syndrome; nfvPPA: non-fluent variant primary progressive aphasia; PSPS: progressive supranuclear palsy syndrome; lvPPA: logopenic variant primary progressive aphasia; bvFTD: behavioral variant frontotemporal dementia; svPPA: semantic variant primary progressive aphasia; sbvFTD: semantic behavioral variant frontotemporal dementia.

To determine the most optimal cut-off point, the sensitivity and specificity values of the DART at each score were calculated when discriminating group pairs, including: All PWD from HC, SCD from OHC, SCD from Other PWD, sbvFTD from OHC, and sbvFTD from Other PWD (Table 3). A DART 10 cut-off score had a 90% sensitivity and 49% specificity (AUC=82%) for discriminating PWD from OHC whereas a DART 9/12 score yielded a 93% sensitivity, and a 67% specificity (AUC=87%) for discriminating SCD from OHC. The greatest accuracy was obtained when differentiating sbvFTD from OHC, with an AUC of 97%, and a DART score of 7/12 showed specificity of 93% and sensitivity of 100%. DART score distinguished the sbvFTD group from all other PWD at a high level of accuracy (AUC 87%); a score of 7/12 discriminated these two groups at 43% sensitivity and 95% specificity (Figure 2).

**Figure 2:**
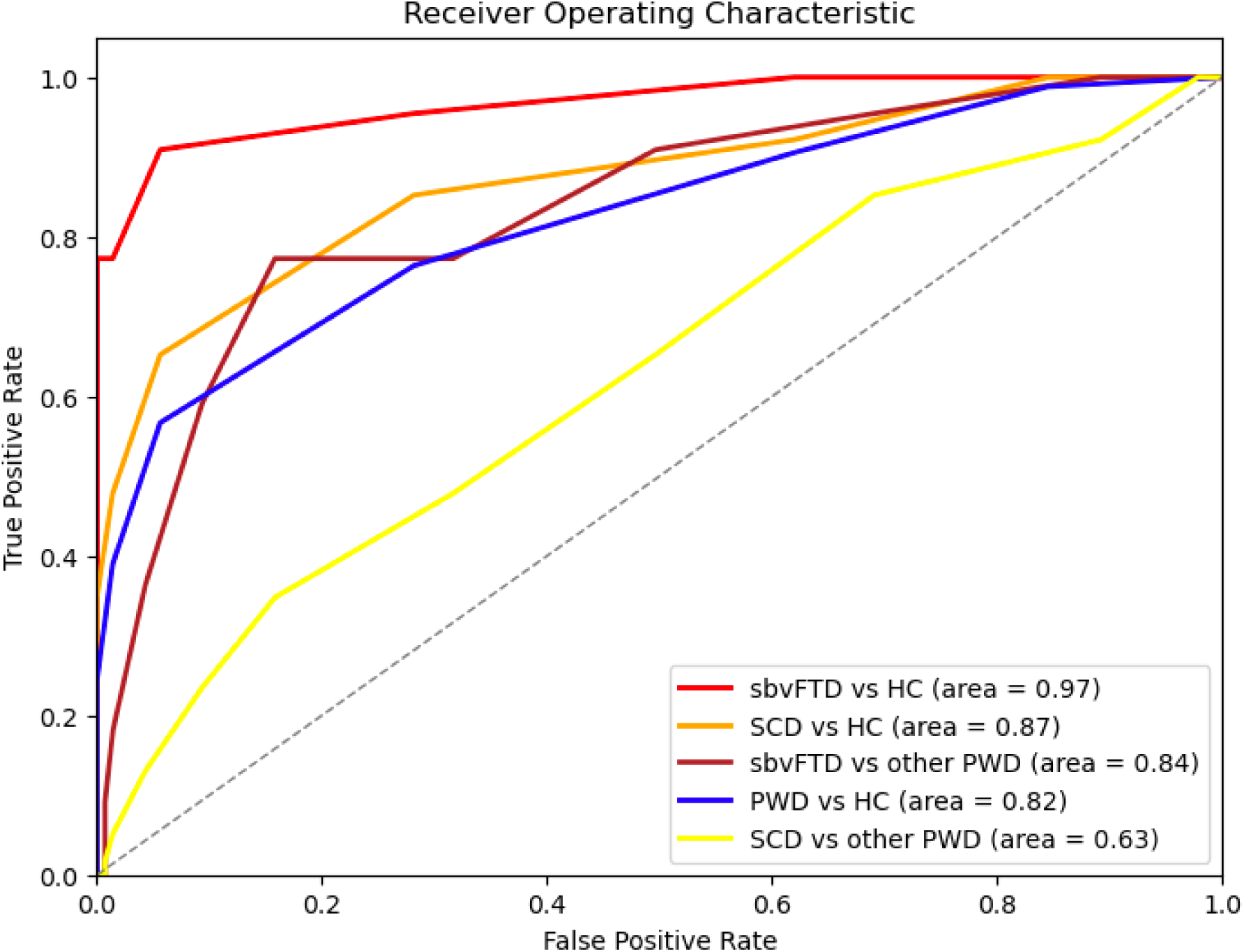
ROC curves for DART total score. PWD: patients with dementia, SCD; social cognition disorder syndromes, including bvFTD, svPPA, and sbvFTD; HC: older healthy controls; sbvFTD: semantic behavioral variant frontotemporal dementia; ROC: receiver operating characteristic; area: area under the curve.

**Table 3:** Sensitivity and specificity of DART throughout score range.

|  | PWD vs. OHCs |  | SCD vs. OHCs |  | sbvFTD vs. OHCs |  | SCD vs. Other PWD |  | sbvFTD vs. Other PWD |  |
| --- | --- | --- | --- | --- | --- | --- | --- | --- | --- | --- |
| DART | Sensitivity | Specificity | Sensitivity | Sensitivity | Sensitivity | Specificity | Specificity | Specificity | Sensitivity | Specificity |
| Score 0 | 0 | 1 | 0 | 0 | 0 | 1 | 0 | 1 | 0 | 1 |
| Score 1 | 0 | 1 | 0 | 0 | 0 | 1 | 0 | 1 | 0 | 1 |
| Score 2 | 1 | 0.23 | 0 | 0 | 0 | 1 | 0 | 0.54 | 0 | 0.86 |
| Score 3 | 1 | 0.24 | 1 | 1 | 1 | 0.84 | 0.66 | 0.55 | 0.66 | 0.87 |
| Score 4 | 1 | 0.24 | 1 | 1 | 1 | 0.86 | 0.75 | 0.56 | 0.66 | 0.88 |
| Score 5 | 1 | 0.25 | 1 | 1 | 1 | 0.89 | 0.71 | 0.57 | 0.57 | 0.90 |
| Score 6 | 1 | 0.27 | 1 | 1 | 1 | 0.92 | 0.67 | 0.58 | 0.50 | 0.93 |
| Score 7 | 1 | 0.29 | 1 | 1 | 1 | 0.93 | 0.64 | 0.60 | 0.43 | 0.95 |
| Score 8 | 0.99 | 0.33 | 0.98 | 0.92 | 0.92 | 0.95 | 0.55 | 0.61 | 0.28 | 0.95 |
| Score 9 | 0.97 | 0.40 | 0.93 | 0.78 | 0.78 | 0.99 | 0.52 | 0.63 | 0.22 | 0.97 |
| Score 10 | 0.90 | 0.49 | 0.79 | 0.42 | 0.42 | 0.98 | 0.50 | 0.71 | 0.18 | 0.98 |
| Score 11 | 0.83 | 0.54 | 0.65 | 0.25 | 0.25 | 1 | 0.46 | 0.62 | 0.15 | 1 |
| Score 12 | 0.79 | 0.79 | 0.59 | 0.20 | 0.20 | 1 | 0.45 | 1 | 0.14 | 1 |

DART: Dynamic Affect Recognition Test; PWD: persons with dementia, SCD; social cognition disorder syndromes, including bvFTD, svPPA, and sbvFTD; OHC: older healthy controls;
sbvFTD:
semantic
behavioral
variant
frontotemporal
dementia

### Brain-Behavior Relationship Underlying DART Performance

VBM analysis of DART total score, corrected by age, sex and TIV, showed that poorer DART performance correlated with focal volume loss in bilateral temporal, frontal, subcortical and cerebellar areas. The highest T-scores were observed in bilateral anterior and posterior insula (R>L), temporal pole, caudate head (L>R), posterior orbital gyrus (R>L), right subcallosal area, right medial orbital gyrus, right superior, middle, inferior and transverse temporal gyrus, right fusiform gyrus, right entorhinal area and left cerebellar crus (pFWE<0.05, T Score>6) (Figure 3, Table 4). To examine whether DART score reflected brain regions that were specific to emotion reading, over and above deficits in confrontation naming or semantic processing, we also analyzed DART controlling for BNT, with the intention to subtract out all regions shared with the BNT performance. Remaining areas specific to the DART appropriately reflected primarily right-sided frontotemporal structures known to correspond to emotion reading. These included the insula, temporal pole, caudate, and superior frontal gyrus pars medialis (pFWE<0.05, T score>6). Correlations with amygdala, hippocampal, and nucleus accumbens regions became non-significant. The region of the brain reflecting the highest T-score in the BNT-corrected analysis of DART was the left cerebellar crus (T=6.63), reflecting the same asymmetry seen in right-side cortical results connected contralaterally through the cerebellar diaschisis.

**Figure 3:**
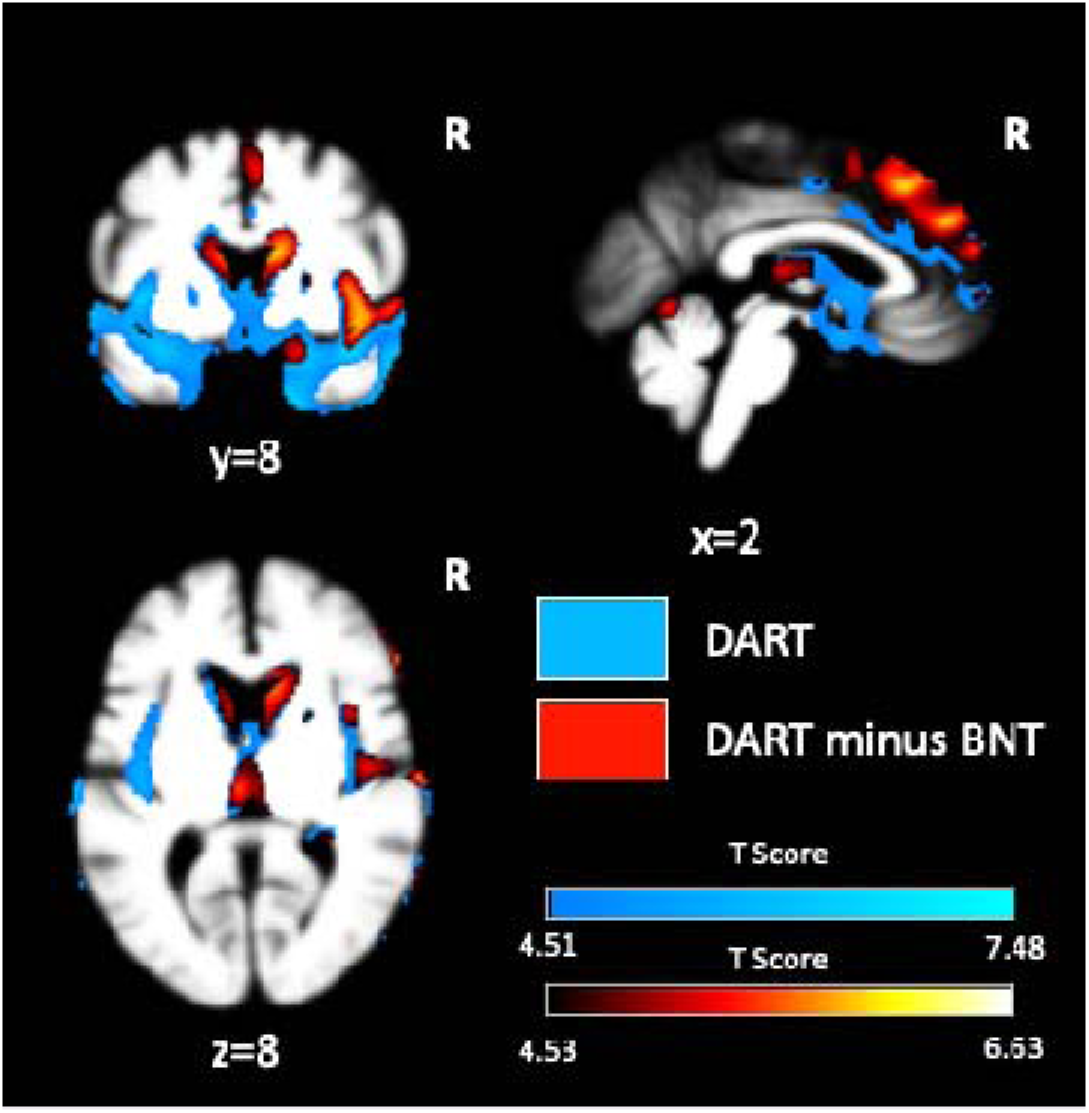
Structural atrophy correlated with poorer DART scores. Voxel-based morphometry analysis of regional brain volume predicted by DART total score (blue), compared to regions reflecting the DART score controlling for Boston Naming Test total score (BNT) (red-yellow). T-maps are corrected for family-wise error at pFWE<0.05. All results are corrected for age, sex, and TIV.

**Table 4:** Structures where volume was significantly predicted by DART total score.

|  | Right Hemisphere |  |  |  | Left hemisphere |  |  |  | After BNT Correction |  |
| --- | --- | --- | --- | --- | --- | --- | --- | --- | --- | --- |
|  | Max T | x | y | z | Max T | x | y | z | Max T | * |
| **Temporal** |  |  |  |  |  |  |  |  |  |  |
| Posterior Insula | 7.47 | 40 | -13 | -4 | 6.37 | -42 | -10 | -6 | 6.39 | R |
| Planum Polare | 7.43 | 40 | -13 | -6 | 6.46 | -43 | -10 | -7 | 6.32 | R |
| Anterior Insula | 7.07 | 42 | 4 | -9 | 6.29 | -39 | 6 | -1 | 6.03 | R |
| Temporal Pole | 6.65 | 40 | 7 | -19 | 6.99 | -48 | 19 | -18 | 5.46 | R |
| Middle Temporal Gyrus | 6.64 | 66 | -45 | 0 | 5.58 | -51 | 1 | -36 | 5.59 | R |
| Fusiform Gyrus | 6.41 | 36 | -21 | -33 | 6.00 | -30 | -30 | -27 | 4.84 | R |
| Inferior Temporal Gyrus | 6.38 | 51 | -9 | -39 | 5.75 | -39 | -7 | -43 | 4.54 | R |
| Superior Temporal Gyrus | 6.19 | 58 | -1 | -13 | 5.98 | -66 | -22 | 6 | 5.83 | R |
| Transverse Temporal Gyrus | 6.17 | 43 | -15 | 0 | 5.72 | -40 | -18 | 4 | 5.47 | R |
| Entorhinal Area | 6.17 | 19 | 3 | -19 | 5.77 | -21 | 0 | -37 | 4.53 | R |
| Amygdala | 5.85 | 16 | -1 | -16 | 5.48 | -16 | -3 | -16 | 4.12 | R |
| Hippocampus | 5.50 | 33 | -36 | -6 | 4.69 | -19 | -13 | -22 | 4.09 | R |
| **Frontal** |  |  |  |  |  |  |  |  |  |  |
| Subcallosal Area | 6.52 | 18 | 3 | -15 | 5.89 | -18 | 1 | -15 | 5.39 | R |
| Medial Orbital Gyrus | 6.45 | 18 | 7 | -21 | 5.96 | -22 | 18 | -24 | 5.24 | R |
| Posterior Orbital Gyrus | 6.07 | 25 | 19 | -22 | 6.04 | -30 | 10 | -21 | 5.79 | R |
| Superior Frontal Gyrus Pars Medialis | 5.80 | 1 | 43 | 28 | 5.81 | -1 | 43 | 27 | 6.01 | R |
| Lateral Orbital Gyrus | 5.70 | 43 | 27 | -16 | 4.68 | -39 | 33 | -18 | 5.37 | R |
| Central operculum | 5.64 | 40 | 7 | 4 | 5.27 | -43 | -9 | 6 | 5.42 | R |
| Frontal operculum | 5.63 | 42 | 9 | 1 | 5.23 | -39 | 9 | 3 | 5.42 | R |
| Supplementer Motor Cortex | 5.58 | 1 | 24 | 40 | 5.53 | 0 | 22 | 40 | 5.87 | R |
| Inferior Frontal Gyrus Pars Orbitalis | 5.43 | 46 | 28 | -15 | 4.79 | -45 | 21 | -13 | 5.07 | R |
| Anterior Cingulate Gyrus | 5.31 | 1 | 37 | 25 | 5.08 | 0 | 37 | 25 | 5.35 | R |
| Inferior Frontal Gyrus Pars Triangularis | 5.18 | 51 | 40 | 3 | 5.15 | -54 | 24 | 1 | 5.89 | R |
| Middle Cingulate Gyrus | 5.08 | 1 | 21 | 24 | 4.90 | 0 | 22 | 22 | 5.12 | R |
| Anterior Orbital Gyrus | 4.35 | 33 | 42 | -16 | 4.23 | -31 | 37 | -16 | 4.66 | R |
| Inferior Frontal Gyrus Pars Opercularis | 4.41 | 57 | 24 | 9 | 4.32 | -52 | 19 | 0 | 5.21 | R |
| **Parietal** |  |  |  |  |  |  |  |  |  |  |
| Angular gyrus | 5.98 | 61 | -52 | 16 | - | -58 | -57 | 24 | 5.23 | R |
| **Subcortical** |  |  |  |  |  |  |  |  |  |  |
| Caudate | 6.03 | 10 | 16 | 10 | 6.45 | -9 | 10 | 13 | 6.00 | R |
| Putamen | 5.44 | 19 | 6 | -12 | 4.95 | -21 | 3 | -10 | 4.66 | R |
| Thalamus | 5.27 | 10 | -6 | 16 | 5.65 | -3 | -19 | 9 | 5.69 | R |
| Accumbens Area | 5.09 | 6 | 9 | -3 | 4.73 | -7 | 18 | -1 | 4.04 | R |
| **Cerebellum** |  |  |  |  |  |  |  |  |  |  |
| Cerebellar Crus | 6.07 | 28 | -85 | -34 | 6.37 | -28 | -84 | -36 | 6.63 | L |
| Cerebellar Vermal Lobules I-V | 4.75 | 1 | -61 | -4 | 4.75 | -3 | -69 | -12 | 5.31 | B |

BNT: Boston Naming Test, R: Right, L: Left, B: Bilateral
\* Predominantly affected hemisphere. Anatomical areas were presented if they surpassed the FWE-corrected significance threshold in either hemisphere (DART= $T > 4.51$ , DART-BNT= $T > 4.53$ ).

## DISCUSSION

This study describes the test characteristics of the Dynamic Affect Recognition Task, a novel, brief, freely-available tool designed for the assessment of emotion recognition through realistic videos, and examines its utility in a large cohort of persons with dementia who are at the earliest stages of disease. The DART shows psychometrically robust characteristics in younger and older neurologically healthy controls, and showed utility for identifying dementia syndromes known to involve social cognition symptoms, with particular sensitivity to individuals with semantic behavioral variant frontotemporal dementia. A DART score of 9/12 was identified as the optimal cutoff to effectively differentiate individuals with social cognitive disorder syndromes (i.e. bvFTD, svPPA, and sbvFTD) from OHCs. Additionally, in PWD performance on the DART was found to correspond focally with brain volume in structures known to mediate emotion reading, particularly within right frontotemporal regions, further validating that the test has construct validity as a measure of emotion recognition in these patients. Overall, our results confirm that DART is an effective tool for differentiating dementia subtypes and identifying patients with focal disruption of the neurologic circuits underlying emotion reading.

### Relevance of the DART for dementia evaluation

In recent years there has been a rapid growth in scientific knowledge derived from clinical research with persons with dementia, which is increasingly relevant to improving day-to- day clinical practice. Recent clarification of the distinct neuropathological entities underlying dementia syndromes underscores the critical significance of precise clinical phenotyping and the necessity for enhanced precision in clinical terminologies pertaining to patients’ behavioral issues (40). Quantitative evaluation of the cognitive changes causing patients’ real-life socioemotional deficits is rapidly becoming essential for comprehensive neuropsychological dementia evaluation. Addressing this need requires the development of cognitive assessment tools that are based in rigorous neurologic science, but are practically useful for widespread implementation. The ultimate benchmark for unraveling symptomatology lies in establishing concordance across caregiver observations, clinician evaluations, and face-to-face objective instruments that uncover the neural mechanisms causing the behavioral problem. Regrettably, traditional behavioral neurology practices disproportionately depend on the first two aspects, often falling short in quantitative testing (40). Numerous endeavors have been undertaken to create tests that bridge this gap; however, the outcomes have not always been successful (4,9,25). This can be attributed to various factors, ranging test designs that are too complex or lengthy for practical administration, to challenges posed by running test validation studies in dementia patients at advanced stages, or using smaller or poorly characterized samples. Our approach to addressing these issues seen in previous studies was to employ a test that was carefully designed for neurologic rigor and practical use, and validating it in a meticulously phenotyped cohort at a very early disease stage, corresponding optimally to the timing when clinical diagnostic evaluation typically occurs. Also, our study benefitted from a larger sample size compared to many other studies in the domain of neuropsychological evaluation of socioemotional functioning (4,9,25). This strategic approach yielded clear results for the DART, with AUC values for diagnostic discrimination exceeding 85%, alongside clear correspondence with targeted brain circuits for emotion reading.

### Cross-cohort applicability of the DART

The adaptation or development of novel tests of social cognition has been identified as a research priority by experts on cross-cultural neuropsychological assessment of dementia throughout North and South America, Europe, and Asia (40–44). The DART has strengths and weaknesses in this regard; positives are the focus on basic emotions with less ambiguity across cultures and the intentional inclusion of racially diverse actors, while a limitation is that the audio is recorded in English. While many studies have found no cross-cultural differences in the recognition of the “six basic emotions” (i.e. happy, surprised, sad, angry, frightened, and disgusted), (45) more nuanced accounts have more recently arisen. For example, a recent cross-cultural study involving 18 centers across 12 countries revealed that while positive emotions were almost universally identified, there were cultural variations in the recognition of negative emotions, (46) which may in part be due to the often-noted ceiling effect for the broad and very easily recognizable positive emotion of happiness. The "attentional limitation hypothesis" has been proposed to elucidate this phenomenon, (47) arguing that in static pictures of emotions, some facial expressions are highly visually similar because they involve only slightly different contractions of the same muscle groups. The DART aims to reduce this confusion by using dynamic, multimodal stimuli and thus adding auditory voice cues and showing the distinct temporal dynamics involved in the expression of each emotion. This has the added benefit of ensuring that test stimuli more closely match real-life emotion expressions, thus will more accurately discriminate patients who are unable to use these additional multimodal cues in real life from those whose day-to-day emotion reading remains normal. Another significant limitation noted in the existing literature is the fact that many well-established emotion reading tasks utilize racially homogenous faces, predominantly of white ethnicity, posing a challenge to cross-cultural adoption for use in non-white groups. (48,49) However, the DART’s design mitigates this concern by involving a racially diverse cast of actors. Furthermore, while actors are speaking in casual English dialogue, thus their idioms may be differentially understood across cultural contexts, these scripts are semantically neutral and comprehension of the spoken content is irrelevant to test performance. This approach was designed to maximize the DART’s adaptability across varying cultural contexts within the English-speaking world; however, it also renders the test highly amenable to translation and refilming in other languages. Versions of the DART in both neutral and Argentinian Spanish have been created and are undergoing validation, and a Mandarin Chinese version is under construction. Results of the validation studies for these non-English versions of the DART will provide valuable insights into the test’s limitations and potential areas for refinement.

Additional confounds to be considered for any neuropsychological measure are education, age, and sex. In our normative sample, spanning healthy controls from the age of 19- 90, we found only very slight impact on DART scores from any of these factors. The influence of education and age on neuropsychological tests of socioemotional cognition, and particularly emotion reading, may generally be less than what is observed in other traditional domains of higher cognition such as memory and visuospatial functioning. While varying outcomes have been observed across studies, one multicenter study with a substantial sample size (n=587) found no significant effect of education on emotion recognition performance. (46) Of note, our normative sample was centered in a Western, Educated, Industrialized, Rich, and Democratic (i.e. WEIRD) sample, (50) therefore our normative scores should be applied with caution in more broadly representative samples until normative datasets from those populations can be collected. Regarding the impact of age, our oldest healthy subgroup aged 70 and above performed slightly lower than younger groups on the DART overall, though the difference was quantitatively small enough that it would not be distinguishable at the individual patient level. A recent meta-analysis has revealed that technical factors such as visual impairments, auditory challenges, or difficulties in maintaining concentration may play a more significant role in the impaired emotion recognition performance of older participants. (51) However, because emotion recognition deficits are one of the earliest symptoms in social cognition disorders, and particularly sbvFTD, (8,13) it is important not to immediately discount lower emotion reading scores as a by-product of age, which could lead to delays in diagnosing their condition. With respect to gender, which has been inadequately separated from biological sex in most neuropsychological research studies thus far but which happened to be fully overlapping in this particular older adult sample, the existing literature currently accommodates two opposing perspectives. The “gender difference hypothesis” suggests that women have better socioemotional decoding skills than men, even if this advantage is relatively small according to meta-analyses. (52) Conversely, the "gender similarity hypothesis" contends that gender disparities often merely reflect experimental biases, and asserts that women and men tend to exhibit considerable similarity across numerous psychological dimensions. (53) Our findings align with the latter proposition. Nevertheless, the influence of cultural gender roles on social cognition, and the early deficits caused by neurodegeneration, requires more exploration. Future studies should explicitly inquire about participants’ gender identity as distinct from their sex at birth or current biological sex, transcending the traditional binary male/female classifications to gain a deeper comprehension of the intricate role sex and gender play in social cognition performance.

### Use of the DART to identify focal brain dysfunction

We support the assertion that a neuropsychological tests used for symptom phenotyping and differential diagnosis in patients with neurodegenerative syndromes should have an empirically established, reasonably focal correspondence with the function of underlying brain circuits, facilitating a neurological interpretation of test performance. Our study found that DART performance corresponded directly with brain regions known to be involved in emotion reading, including predominantly right anterior and posterior insula, temporal pole, caudate, superior frontal gyrus pars medialis and supplementary motor cortex, as well as the left cerebellar crus. Meta-analyses focusing on the anatomical underpinnings of emotion recognition, coupled with more specialized investigations within neurodegenerative disease samples, have illuminated the roles of various visual, limbic, temporoparietal, and prefrontal regions, alongside basal ganglia and cerebellum. (5,9,54–56) Particularly after the adjustment using BNT, the predominantly right-sided nature of our DART results was consistent with evidence from a recent meta-analysis arguing that right sided structures receive more sensory input relevant to emotional experience than the left. (57) Among those right-lateralized cortical structures, the most impacted regions encompassed the insula, recognized by several authors as a central contributor to multimodal emotion processing of inputs ranging from basic sensory stimuli to higher-order cognition; (13,58–61) the temporal pole, hailed as the hub of emotional semantics; (5,57,62) the superior, medial, and inferior temporal gyri, which have long been associated with both auditory (i.e. voice prosody) and visual (face and body viewing) aspects of person perception and affect sharing; (56) the orbitofrontal gyrus, involved in tagging socioemotional concepts with a hedonic evaluation; (10,63) the medial superior frontal gyrus, inferior frontal gyrus and supplementary motor area, identified as cortical control, regulation and production areas of emotion; (64–67) and lastly the anterior cingulate, a central hub of the salience network, directing attentional resources towards emotional stimuli. (13) The caudate and thalamus were the predominantly affected subcortical areas, which is not unexpected considering their critical role in socioemotional cognition related networks and emotion regulation. (13,56,64,65) A particularly noteworthy finding was the strong correspondence with volume in the cerebellum, underscoring its important but as yet underexplored role in social cognition. (68)

### Limitations and Conclusions

This study was designed to assess the utility of the DART in patients with various neurodegenerative disease syndromes, some of which are relatively rare. Consequently, certain group sizes were comparatively small. However, it should be noted that to our knowledge, this is still one of the largest socioemotional test validation studies in this population where patients are at a very early stage with very focal atrophy, and have concurrent structural MRI scans. Also, because many of these subtypes are encountered relatively infrequently, our study design may yet provide adequate information for discriminating them on a practical basis. Further data collection of both healthy controls and patients in non-WEIRD samples is also needed, as we have detailed above. Lack of genetic and pathological confirmation of diagnosis could be considered another limitation of the study, though most cases in these dementia subtypes are of sporadic origin. To mitigate any impact of lack of pathological confirmation in our sample, patients received diagnoses from a highly experienced multidisciplinary team, following comprehensive cognitive and neurological evaluations. Moreover, each patient had supporting neuroimaging evidence, substantially bolstering diagnostic accuracy. In this validation study, normative values and cut-off scores were provided to differentiate persons with dementia, particularly those with social cognition disorder syndromes, from healthy controls, facilitating application of the DART for diagnostic use. However, similarity in the range of DART scores between many pairs of syndromes meant that this measure was not precise enough to derive reliable cut-off scores to differentiate specific dementia subtypes from one another. Finally, it will be important for future studies to investigate the mechanistic relationship between emotion reading and other cognitive functions across diverse neurodegenerative syndromes.

In conclusion, the DART is a brief, psychometrically robust video-based test of emotion reading that (i) is designed to be practically useful in realistic clinical research and clinical settings, (ii) effectively reveals emotion identification impairments in PWD, (iii) shows specificity for identifying PWD exhibiting real-life social cognition deficits (i.e. bvFTD, svPPA, sbvFTD), (iv) corresponds to the expected structural anatomy of emotion reading, and (v) is freely available to clinicians and researchers, either via the UCSF TabCAT platform available on the Apple store, or via direct contact with the authors. Because socioemotional symptoms are not only of central clinical importance to patients and family members, but also have significant value for differential diagnosis and prognostication in persons with neurodegenerative disease, it is important that the field continues to pivot towards routine use of face-to-face neuropsychological tests like the DART in diagnostic and staging evaluations.

## Acknowledgements

We are most thankful to all participants, their family members and the volunteers that participated in this research, and the project’s research coordinators who helped with data collection, quality, and management: Patrick Callahan, Bailey McEachen, Faatimah Syed, Myrthe Rijpma, Rea Antoniou, and Cesar Hernandez. We also are so grateful to our actors who generously shared their time to be filmed for the DART videos. VBM analyses were performed using the open-source UCSF Brainsight system, developed at the UCSF Memory and Aging Center by Katherine P. Rankin, Cosmo Mielke, Paul Sukhanov, and Rain Simons, and powered by the VLSM script written by Stephen M. Wilson, with funding from the Rainwater Charitable Foundation and the UCSF Chancellor’s Fund for Precision Medicine.

